# An interpretable and explainable neural network to classify sports-related cardiac arrhythmias in professional football athletes

**DOI:** 10.64898/2026.02.23.26346628

**Authors:** Erik Vanegas Müller, Mirae Harford, Liang He, Abhirup Banerjee, Paul Leeson, Mauricio Villarroel

## Abstract

Sudden cardiac death risk is 2-3-fold higher in athletes than in non-athletes. We classify sports-related cardiac arrhythmias using a novel explainability framework comprising data analysis, model interpretability, post-hoc visualisation, and systematic assessment. Two neural networks—one with interpretable sinc convolution and one with standard convolution—were trained on general-population ECGs (PhysioNet, n=88,253, 30 arrhythmias, three continents) and tested on professional footballers (PF12RED, n=161) via domain adaptation for normal sinus rhythm (NSR), sinus bradycardia (SB), incomplete right bundle branch block (IRBBB), and T-wave inversion (TWI).

Sinc convolution achieved superior NSR detection (AUROC 0.75 vs 0.70), whilst standard convolution excelled at SB (0.74 vs 0.73), IRBBB (0.66 vs 0.58), and TWI (0.59 vs 0.54). Gradient-weighted Class Activation Mapping revealed that sinc models focus on physiologically relevant ECG segments (the PR interval for NSR/SB and the T wave for TWI). We hypothesise that sinc convolution better captures periodic rhythms but struggles with complex morphological patterns, suggesting architectural choice should align with underlying cardiac pathophysiology.

**Graphical abstract:** Abbreviations: AI, artificial intelligence; AUPRC, area under the precision-recall curve; AUROC, area under the receiver operating characteristic curve; Conv, convolution; ECG, electrocardiogram; Grad-CAM, gradient-weighted class activation mapping; IAVB, first-degree atrioventricular block; IRBBB, incomplete right bundle branch block; LAD, left axis deviation; LBBB, left bundle branch block; LVH, left ventricular hypertrophy; NSR, normal sinus rhythm; QT, QT interval; RAD, right axis deviation; RBBB, right bundle branch block; RVH, right ventricular hypertrophy; SA, sinus arrhythmia; SB, sinus bradycardia; TWI, T-wave inversion; xAI, explainable artificial intelligence.

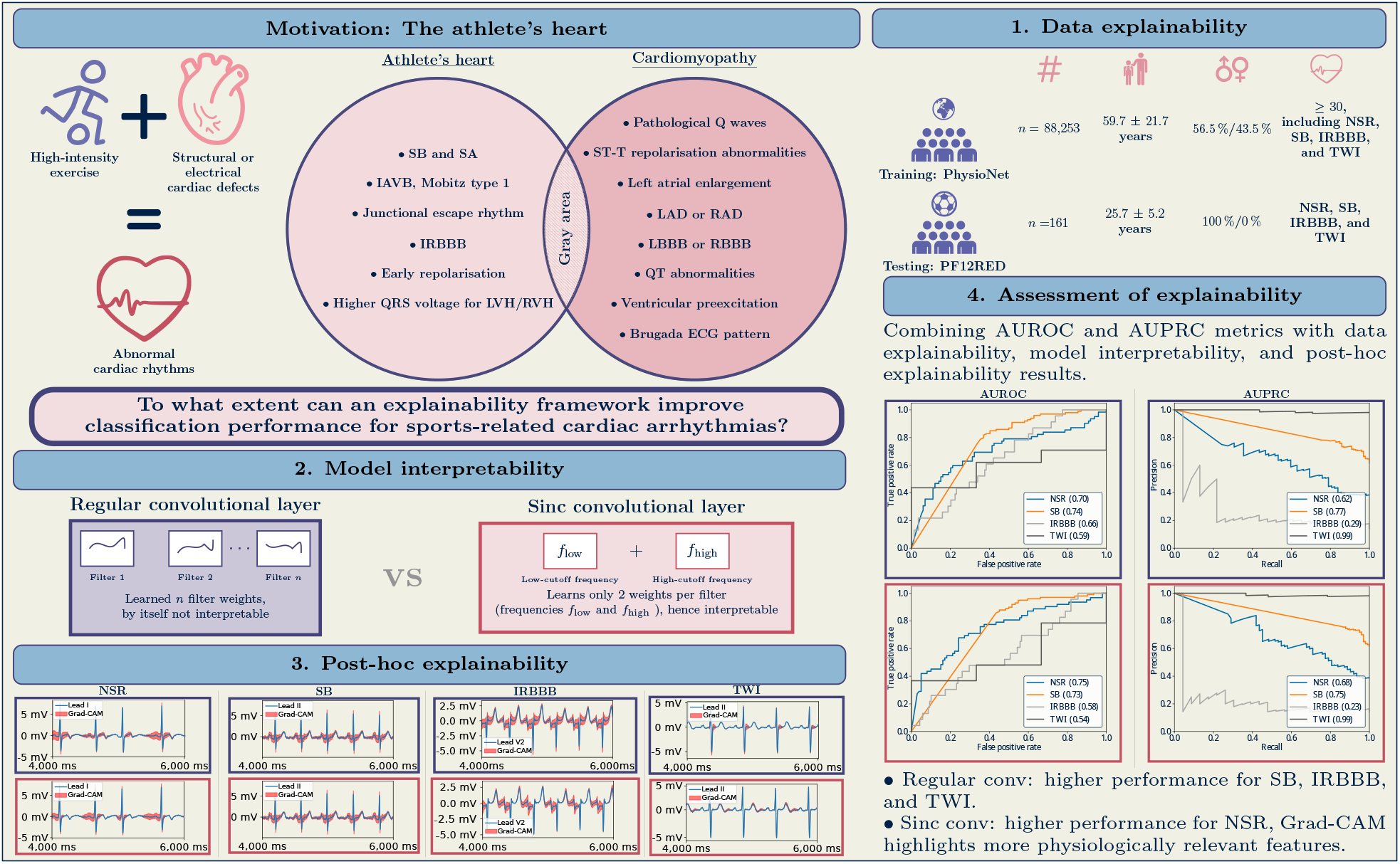

## Introduction

Sudden cardiac death (SCD) is the unexpected death due to a cardiovascular cause, occurring within one hour of witnessed or 24 hours of unwitnessed cardiac arrest from symptom onset. Despite the well-known benefits of exercise, the risk of SCD for athletes doubles during physical activity and is higher while exercising than in non-athletes ^1-4^. Exercise-induced cardiac adaptations can evolve into an ambiguous area between pathological and physiological adaptations (see Appendix A). Undiagnosed heart diseases, including inherited or acquired cardiomyopathies (disease of the heart muscle) or channelopathies (dysfunction of ion channels), can lead to life-threatening arrhythmias ^5^.

Huttin et al. ^6^ studied exercise-induced Electrocardiogram (ECG) changes in 2,484 French professional football players. The serial evaluations performed between 2005 and 2015 include 6,247 12-lead ECGs. Sinus bradycardia (SB) at 98%, first-degree atrioventricular block at 31%, and short PR Interval at 80% (including incomplete right bundle branch block (IRBB)) are the main ECG-related exercise-induced cardiac adaptations observed in the cohort.

Professional athletes undergo pre-participation screenings to risk-stratify for adverse arrhythmias against physiological cardiac adaptations. The ECG provides a non-invasive, diagnostic screening modality. The ECG signal represents cardiac depolarisation and repolarisation, and the number and position of leads determine the coverage of the heart’s electrical activity. Noise, such as the body movement artefacts or power line interference, can superimpose the ECG signal and complicate the athlete’s diagnosis ^7^.

Artificial intelligence (AI) methods can extract complex ECG morphologies, classify arrhythmias, and predict outcomes of cardiovascular diseases ^8,9^. However, inferring interpretability and explainability of AI’s outcomes is limited by AI’s black-box nature. Interpretable machine learning (iML) and explainable AI (xAI) are approaches to assessing the outcomes of AI algorithms. We define interpretability as the process of understanding an AI model’s underlying workings, and explainability as the process of the model’s decision-making mechanisms ^10^.

Goettling et al. ^11^ analysed short- and long-term ECG features independently by two different AI models. An ensemble combines both models and perturbation analysis outputs. The novelty does not lie in the explainability method per se, but in the deep learning architecture that analyses short- and long-term features separately. Jang et al. ^12^ developed a generative counterfactual xAI. The framework explores how the AI responds to generated counterfactual ECGs, analysing to what extent modifications in ECG morphologies affect the AI model’s predictive value. While these articles use novel interpretable architectures or single explainability methods, they lack a methodological framework that integrates different explainability axes —such as data explainability, model interpretability, post-hoc explainability, and explainability assessment— into a pipeline to enhance model robustness and trustworthiness ^10^.

Open-source sports-related ECGs are scarce. Domain adaptation studies the transfer of knowledge from a general source, such as a large, heterogeneous ECG dataset, to a specific source, such as a scarce dataset of athletes’ ECGs ^13^. Existing data-rich cardiac electrophysiological signals from general-population hospital visits can aid the design of new AI algorithms for athletes through domain adaptation.

We propose an xAI pipeline for classifying sports-related cardiac rhythms and arrhythmias in young professional footballers. We develop two neural networks: one with an interpretable architecture (using a sinc convolutional layer) and one with standard architecture (using a regular convolutional layer). The sinc convolution constrains the neural network processes physiologically relevant frequencies (e.g., the P-wave, QRS-complex, or T-wave) in the ECG signal. The regular convolutional layers, on the other hand, learn patterns that may not correspond to interpretable signal characteristics.

We train the networks on hospital ECG data from the general population (PhysioNet) and test them on sports-related data (PF12RED). We examine the data imbalance in both datasets. We use Gradient-weighted Class Activation Mapping (Grad-CAM) to highlight important ECG regions and focus on differences between the two networks. This approach forms an explainability framework by integrating data explainability (data imbalance in PhysioNet and PF12RED), model interpretability (sinc convolution layer), and post-hoc explainability (Grad-CAM). We expect this explainability framework to provide a more refined and robust assessment of explainability than single methods, particularly given the ambiguity between physiological and pathological exercise-induced cardiac adaptations.

## Methods

### Datasets

We use the following two datasets:

**PhysioNet Challenge 21** ^14^ comprises 88,253 ECGs representing 30 arrhythmias from the general population across seven institutions and three continents, recorded in 3-lead and 12-lead formats (between 257Hz and 1,000Hz, and a time duration from 10s to 1,800s).

**Pro-Football 12-lead Resting Electrocardiogram Database (PF12RED)** ^15^ comprises 161 12-lead ECGs from Spanish La Liga players, collected over five seasons (2018–2022) at rest (500Hz, 10s duration), interpreted according to the 2017 International Criteria for Electrocardiogram Interpretation in Athletes ^16^.

Table 1 summarises dataset characteristics. Key differences include mean age (PhysioNet: 59.7 years; PF12RED: 25.7 years) and arrhythmia distributions: IRBBB (1.4% vs 14.6%) and TWI (3.1% vs 98.1%). The PhysioNet dataset includes ECGs from China (63%), Europe and Russia (25%), and the USA (12%). The PF12RED dataset was collected in Spain and includes players of African, Latin American, Caucasian, and Asian descent.

**Table 1.**
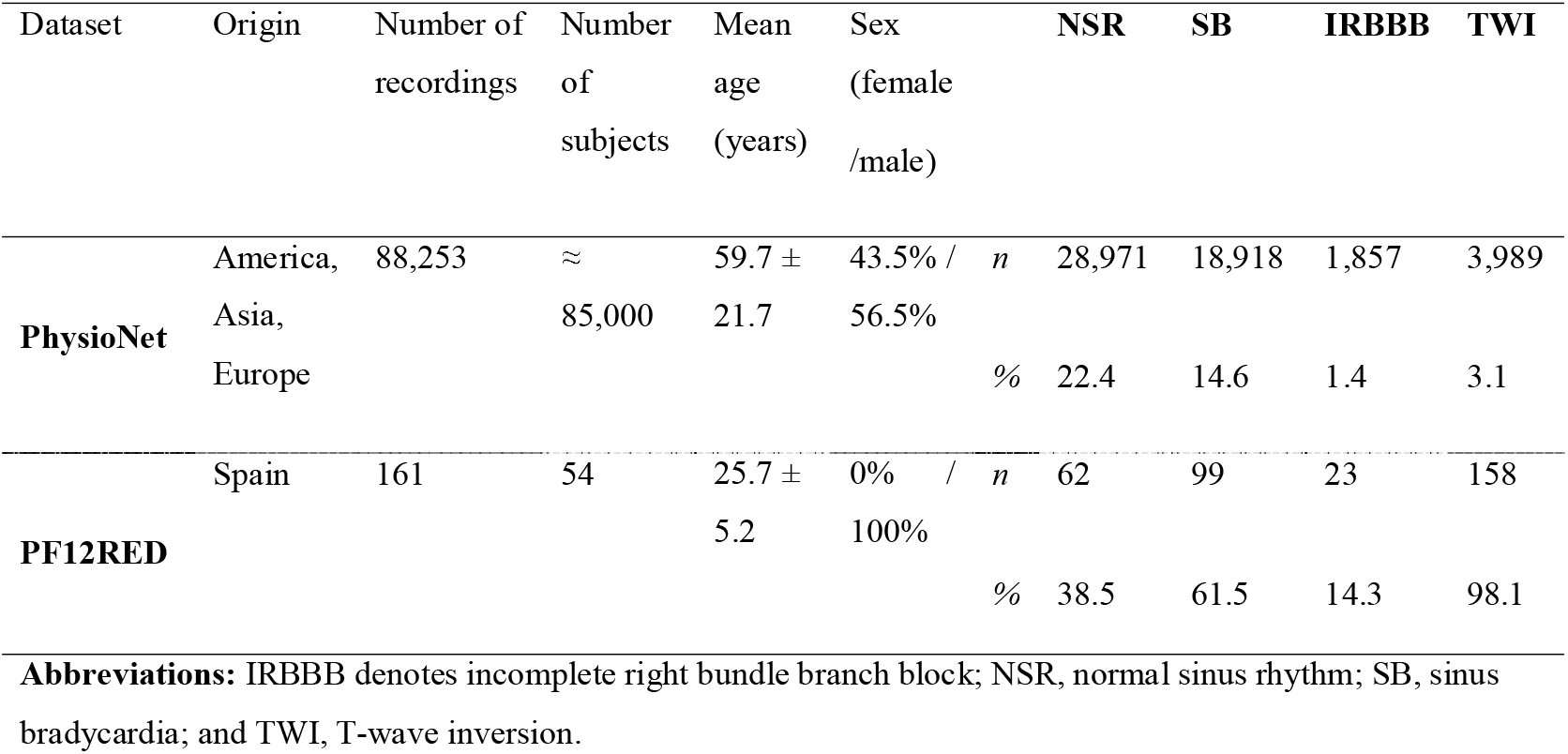
General overview and distribution of arrhythmias in the PhysioNet dataset ^17^ and PF12RED dataset ^15^. The general overview includes the number of recordings, patients, mean age, and sex distribution. The distribution of arrhythmias only shows the cardiac rhythms present in both the PhysioNet and PF12RED datasets. For a complete overview, such as sampling frequency, mean recording duration, labelling procedure, and ECG evaluation criteria, consult Table S3 in the appendix.

### Deep neural block

Figure 1A outlines the workflow. We use a residual neural network with multi-head attention (17) (see Fig. 2B) consisting of two heads (regular or sinc convolution) and a ResNet tail. Both heads use 1×15 filters with a stride of 2, producing 256 channels normalised by batch normalisation and leaky ReLU activation. Output feeds into five subsequent residual blocks (part of the ResNet tail), each containing nine convolution layers with a stride of 2.

**Figure 1.**
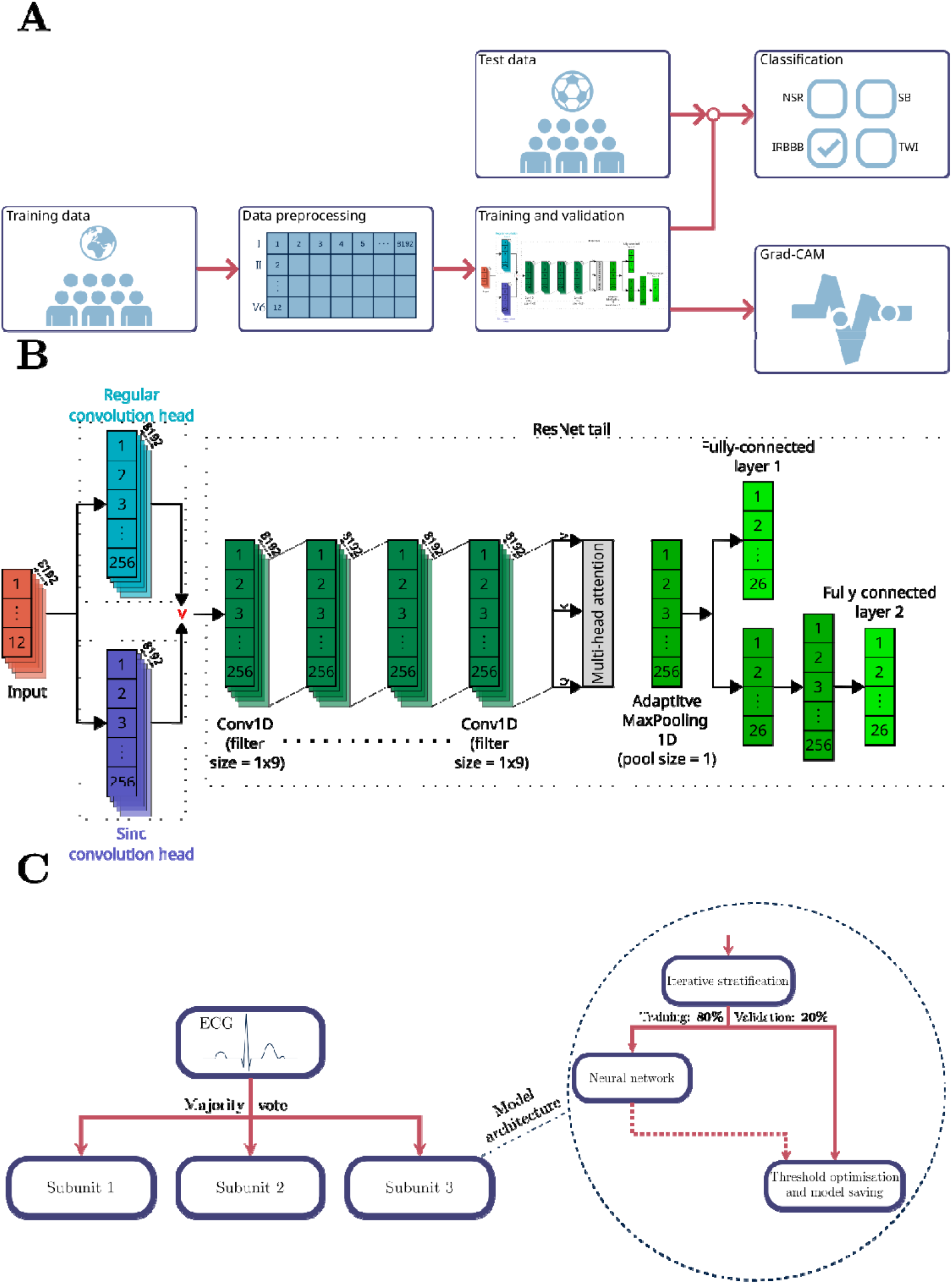
High-level workflow diagram and neural network architecture. **A)** We input preprocessed ECGs from the general population into the neural network. After training and validation, we test the model on a sports-related ECG dataset. The ECG classification is one of the two neural network outputs, the other being Grad-CAM. In this example, the ECG is classified as an IRBBB. We calculate the AUROC, AUPRC, and F1-score to assess the model’s performance. The second output is the Grad-CAM algorithm, which identifies and highlights the most critical regions of the ECG that contribute to the model’s classification decisions. Notice that the Grad-CAM algorithm outputs heatmaps regarding the ECGs used for training and validation. **B)** The neural network architecture. The input goes through either the regular convolution head or the sinc convolution head. The ResNet tail has two outputs: the fully connected layer is forwarded into a BCE loss function. The second output, fully connected to layer 2, is generated by an additional small neural network without backpropagation and is sent to a sparsity and challenge loss function. This design allows the model to focus on the primary task (binary classification) while also optimising for additional tasks or behaviours (sparsity and challenge) that can improve performance and generalisation. **C)** The final model is an ensemble of three subunits, each generating binary indicators for every arrhythmia class. Each subunit comprises the neural network architecture described in B). The final classification is determined through majority voting. For the training-validation split (80:20), we iteratively stratified the PhysioNet dataset to ensure a balanced distribution of arrhythmia classes. The class-specific thresholds are optimised using differential evolution genetic algorithms. AUPRC denotes area under the precision-recall curve; AUROC, area under the receiver operating characteristic curve; BCE, binary cross entropy; Grad-CAM, gradient-weighted class activation mapping; IRBBB, incomplete right bundle branch block; and ResNet, residual neural network.

**Figure 2.**
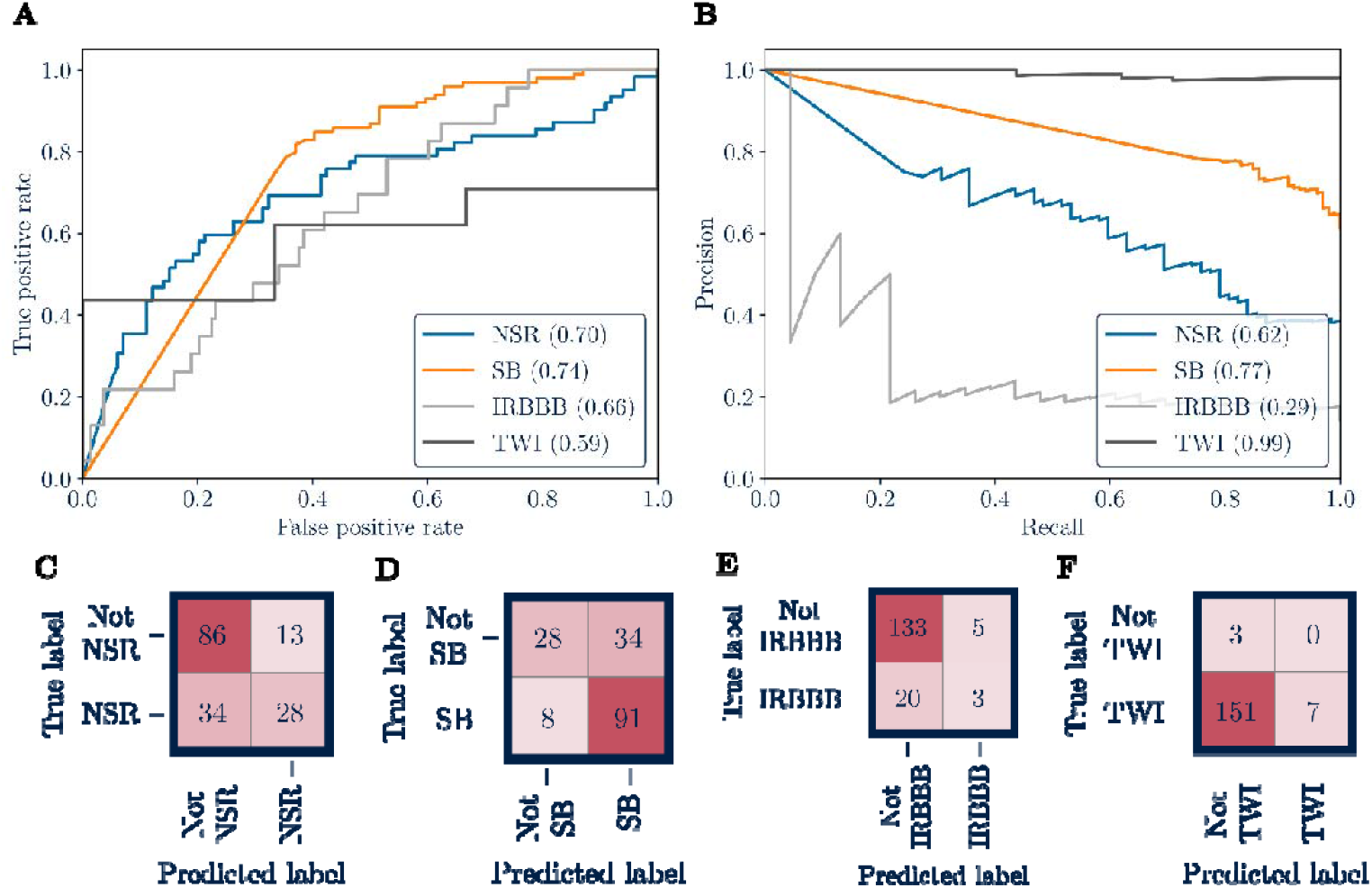
Results for the regular convolution neural network. Plots for **A)** AUROC and **B)** AUPRC, and confusion matrices for **C)** NSR, **D)** SB, **E)** IRBBB, and **F)** TWI. The F1-scores are 0.54 (NSR), 0.81 (SB), 0.19 (IRBBB), and 0.08 (TWI).

### Model interpretability: The sinc convolution layer

Regular convolutional layers learn arbitrary feature patterns without inherent interpretability by applying a mathematical convolution operation to the ECG signal and the learned filter patterns. The learned pattern may not correspond to physiologically meaningful characteristics. In contrast, the sinc convolution layer (see Fig. 2B) constrains the first layer to learn “low- and high-cut-off frequencies of band-pass filters” ^18^. This constraint enhances model interpretability. The interpretable sinc convolution ensures that the network processes ECG signals in a physiologically meaningful way, focusing on specific frequency ranges. The learned low- and high-cut-off frequencies are then passed to subsequent regular convolutional layers. Examples of ECG frequency ranges are 5–30Hz for the P-wave, 8–50Hz for the QRS complex, and 0–10Hz for the T-wave ^19^.

Higher frequencies above 70Hz characterise abnormal conduction ^19^. For instance, learned frequencies above 70Hz would indicate abnormal conduction, which the subsequent convolutional layers could then classify into specific arrhythmia types through pattern recognition. The sinc convolutional layer is explained in greater detail in Appendix D.

### Training strategy

The training approach uses iterative stratification with an 80/20 training-validation split for 30 epochs. The final classification model is an ensemble of three subunits (see Fig. 1C). Each subunit comprises the deep neural network shown in Fig. 1B. After training, class-specific classification thresholds are optimised using differential evolution ^20^, a stochastic search algorithm that does not require gradient information. The model ensemble uses majority voting for arrhythmia classification. The model ensemble of the three subunits uses majority voting for arrhythmia classification.

### Post-hoc explainability: Grad-CAM

Grad-CAM is a visualisation technique that generates explanations as heatmaps of a classification’s critical regions. Grad-CAM is highly class-discriminative ^21^. Explanations regarding a particular cardiac rhythm, such as SB, only highlight morphological regions related to SB but not to TWI or IRBBB. We include an explanation of how Grad-CAM works in Appendix G.

### Evaluation metrics

An appropriate choice of performance metrics is necessary when data are imbalanced (i.e., class sizes are disproportionately large compared to others), especially in the healthcare domain, where model selection (e.g., sinc vs. regular convolutional layers) and performance comparisons depend on these metrics ^22^. We evaluate model performance using the area under the receiver operating characteristic (AUROC), the area under the precision-recall curve (AUPRC), and the F1-score. AUPRC and F1-score are particularly valuable for our imbalanced arrhythmia data, as they focus on true positive detection of target arrhythmias. While AUROC can be influenced by class imbalance, potentially showing optimistically high values due to the abundance of true negatives, it remains valuable as it evaluates discrimination ability across all possible classification thresholds.

## Results

This section presents the results of the neural networks with regular and sinc convolution heads trained on PhysioNet and tested on PF12RED.

Figure 2 shows the results of the regular convolution head. All four cardiac rhythms result in an AUROC above 0.5, with NSR and SB scoring highest (AUROC 0.74) and TWI lowest (0.59). AUPRC ranged between 0.29 (IRBBB) and 0.99 (TWI). F1-score is highest for SB (0.81) and lowest for TWI (0.08). The most common false positives are complete right bundle branch block (CRBBB) (n=34), First-degree atrioventricular block (IAVB) (n=29), right axis deviation (RAD) (n=19), sinus arrhythmia (SA) (n=15), and T-wave abnormality (TAB) (n=12).

Figure 3 presents the results of the sinc convolution head. The network reaches AUROC >0.5 across all rhythms, with the lowest for TWI (0.54) and the highest for NSR (0.75). AUPRC ranges between 0.23 (IRBBB) and 0.99 (TWI). TWI has the lowest F1-score (0.05), SB the highest (0.81). Most common false positives are IAVB (n=35), CRBBB (n=27), SA (n=20), left axis deviation (n=18), RAD (n=11), and TAB (n=10). Comparing F1-scores between models, performance is similar for NSR and SB, whilst regular convolution achieves better scores for IRBBB and TWI.

**Figure 3.**
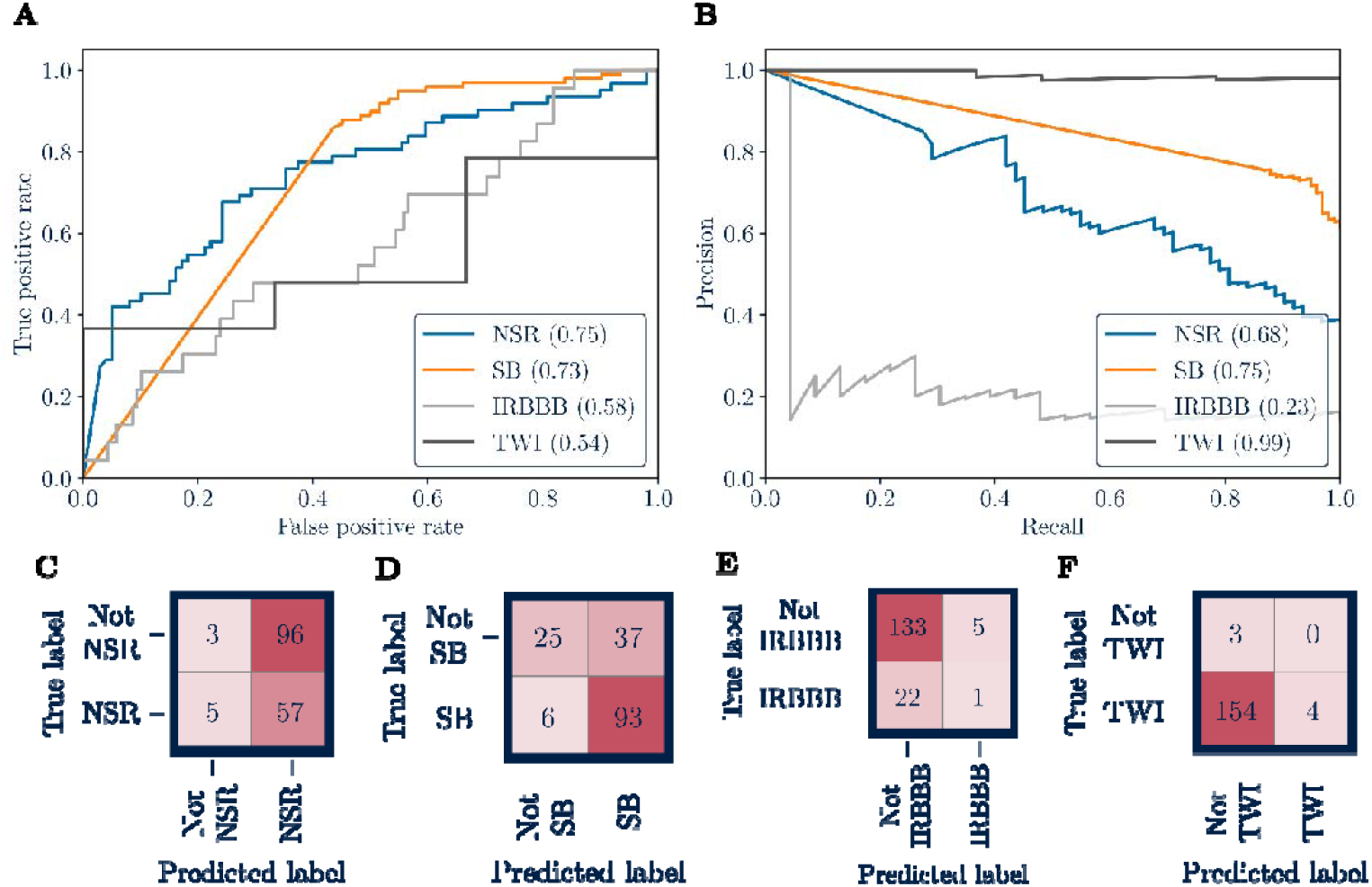
Results for the sinc convolution neural network. Plots for **A)** AUROC and **B)** AUPRC, and confusion matrices for **C)** NSR, **D)** SB, **E)** IRBBB, and **F)** TWI. The F1-scores are 0.53 (NSR), 0.81 (SB), 0.07 (IRBBB), and 0.05 (TWI).

In Grad-CAM, the sinc convolution network highlights zero-padding as important in Figs. 4B, D, F, and H, whilst this behaviour is only observed in Figs. 4C and E for regular convolution. For NSR, Fig. 4B primarily highlights the PR interval, whilst Fig. 4A additionally emphasises the T-wave. SB visualisations in Figs. 4C and D and IRBBB in Figs. 4E and F demonstrate similar activation patterns, although IRBBB showed the least consistent Grad-CAM patterns. For TWI, Fig. 4H shows Grad-CAM focusing on the TP segment, whereas Fig. 4G highlights the ST segment.

**Figure 4.**
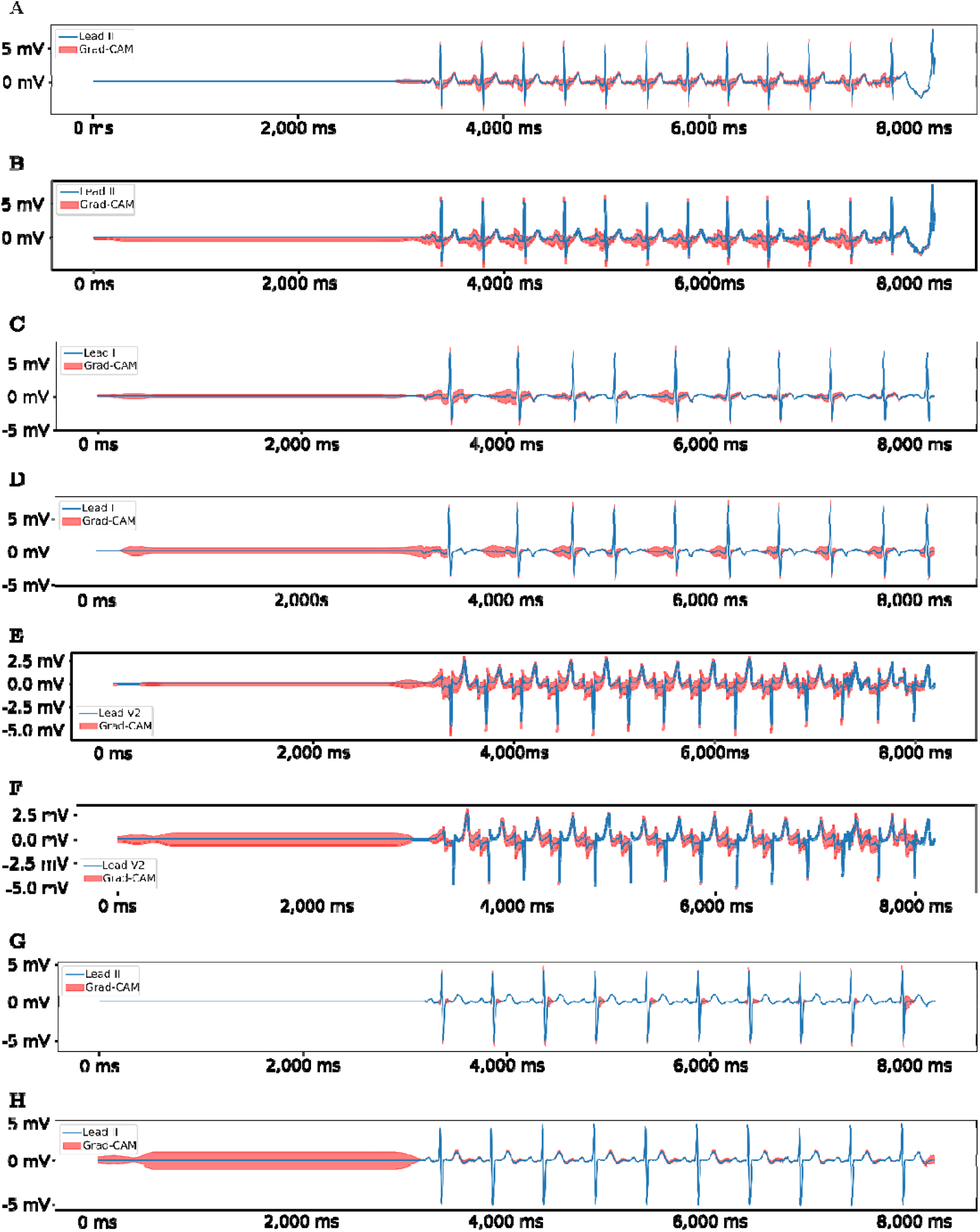
Grad-CAM results for the regular and sinc convolution network regarding NSR, SB, IRBBB, and TWI. For the NSR Grad-CAM results for **A)** regular and **B)** sinc convolution neural network. The patient is a female in their 50s. SB Grad-CAM results for regular **C)** and sinc **D)** convolutional neural network. The patient is a female in their 60s. We chose lead I because it had less noise than the standard lead II. The ECG header file also noted premature atrial contraction, Q-wave abnormality, and T-wave abnormality. IRBBB Grad-CAM results for regular **E)** and sinc **F)** convolutional neural network. The patient is a male and in their 60s. The ECG header file also noted premature atrial contraction, non-specific ST-T abnormality, and left atrial enlargement. TWI Grad-CAM results for regular **G)** and sinc **H)** convolutional neural network. The patient is female and in their 60s. The ECG header file also noted left ventricular hypertrophy and a pacemaker rhythm. Note that the TWI here is biphasic, initiating with a positive and ending with a negative deflection. Because the T-wave ends with a negative deflection, it is considered a TWI.

Both neural networks achieve satisfactory classification (i.e., AUROC > 0.5) for all four cardiac rhythms, with sinc convolution performing better for NSR (AUROC 0.75 vs 0.70) but worse for IRBBB and TWI. F1-scores are comparable for NSR (0.54 vs 0.53) and SB (0.81 vs 0.81), while regular convolution performs better for IRBBB (0.19 vs 0.07) and TWI (0.08 vs 0.05). Grad-CAM visualisations reveal different activation patterns between architectures, particularly in zero-padding attention and ECG segment focus.

## Discussion

SB, IRBBB, and TWI are common exercise-induced cardiac adaptations in football players (see supplementary Appendix A and Fig. S1–S3). Given the scarcity of open-source sports-related ECGs, we use domain adaptation to train neural networks on general-population cardiac arrhythmias (PhysioNet Challenge 21) and test on athlete data (PF12RED). Baseline validation on PhysioNet achieves challenge scores of 0.57-0.59 (30 epochs), comparable to Nejedly et al.’s 0.63 (50 epochs) ^23^. Given the epoch difference, we consider the neural network validated for PhysioNet training/validation and PF12RED testing.

The sinc convolution network performs best at identifying NSR (0.75), followed by SB (0.73). The regular convolution network identifies SB best (0.74) and NSR second (0.70). As shown in Figs. 3C–F, the sinc convolution head often falsely predicted NSR (n=96) and SB (n=37). The regular network showed similar SB misclassification patterns, but differed for NSR, predominantly producing false negatives rather than false positives. We hypothesise that data imbalance influences misclassification rates, as the NSR and SB proportions differ substantially between PhysioNet (22.4% and 14.6%) and PF12RED (38.5% and 61.5%).

SB is considered normal in highly trained athletes due to exercise-induced cardiac adaptations increasing vagal tone, leading to a lower resting heart rate ^24^. The NSR’s average heart rate in Spanish football players is 55 bpm, which is considered normal when asymptomatic. The analysis of NSR and SB behaviour demonstrates both the challenge and feasibility of domain adaptation: whilst transferring knowledge from the general population to athlete ECGs requires accounting for differences in class distributions and population-dependent clinical interpretation.

Grad-CAM visualisations in Figs. 4A-H show essential differences between NSR and SB. The regular convolution focuses on both PR interval and the T-wave. The sinc convolution is more selective, focusing predominantly on the PR interval. For sinus rhythm (irrespective of NSR or SB), pace-making impulses must arise from the sino-atrial node (generating P-waves), PR intervals must be constant (120-200ms), and QRS complexes must be narrow (<100ms) ^25^. Hence, sinc convolution demonstrates more precise activation mapping in Grad-CAM. The enhanced interpretability and band-pass filter behaviour force the network to focus on more physiologically relevant features for NSR and SB identification.

IRBBB has two diagnostic morphologies: RSR’ patterns in V1-V3 (“M”-shaped QRS, Fig. S4) and wide, slurred S waves in lateral leads I, aVL, and V5-V6. Regular convolution outperforms sinc convolution (AUROC: 0.66 vs 0.58; F1-score: 0.19 vs 0.07) due to IRBBB’s morphological rather than frequency-driven nature. Low AUPRC values indicate that both networks struggle with class imbalance, since IRBBB represents only 1.4% of the training data (PhysioNet) but 14.3% of the test data (PF12RED). Most of the IRBBB Grad-CAM results did not show a clear pattern for either the regular or sinc convolution neural network. Figure 4E and F show an example where this is not the case. However, the Grad-CAM highlighting is not as specific to the just-described diagnostic morphology of the IRBBB.

The explainability assessment of the confusion matrices highlights a critical consideration: the target population determines the optimal trade-off between false-positive and false-negative rates. Clinically, minimising false negatives is essential as missing SCD-related conditions could be fatal. For players and clubs, reducing false positives is equally relevant, as both networks misclassified many healthy athletes. The optimal threshold depends on whether priority is placed on comprehensive abnormality detection or on minimising unnecessary exclusions from athletic participation.Common false positives for both networks were CRBBB, IAVB, and RAD, suggesting morphological similarities between these arrhythmias and target conditions in athlete ECGs.

Both networks, especially sinc convolution, highlight zero-padding in Grad-CAM as critical. This highlighting is incorrect, since zero-padding is a pre-processing technique that lacks physical meaning and is not present in real ECGs. We hypothesise that zero-padding degrades sinc convolution performance for SB, IRBBB, and TWI. Despite this, the sinc network demonstrates superior interpretability by focusing on physiologically relevant ECG segments (the PR interval for NSR/SB and the T-wave for TWI).

Summarising, the sinc convolution neural network has the best NSR detection performance (AUROC 0.75 vs 0.70), whilst the regular convolution neural network achieves better SB (AUROC 0.74 vs 0.73), IRBBB (AUROC 0.58 vs 0.66) and TWI (AUROC 0.54 vs 0.59) results. We consider that the sinc model’s frequency-selection properties benefit rhythm-based condition detection, such as NSR, whilst regular convolution’s broader feature-extraction capabilities better capture morphological changes characteristic of IRBBB and TWI.

## Limitations

A central challenge is the severe class imbalance and distribution shift between training and test datasets. The underrepresentation of IRBBB and TWI during training leads to significant misclassification, while dataset specificity from training on the general population and testing on athlete ECGs creates generalisability challenges. Both neural networks struggle to fully adapt to these differences, suggesting that the domain adaptation approach, whilst achieving satisfactory performance, would benefit from larger sports-related training datasets.

Methodological limitations include zero-padding effects, as Grad-CAM highlights how pre-processing techniques can mislead neural networks. Dynamic time warping offers a promising alternative. Additionally, Grad-CAM’s local explainability nature limits interpretation; future work should apply global attention visualisation techniques.

Our study does not adequately address the clinical risk differential among various arrhythmias, as not all cardiac abnormalities are clinically equivalent. For instance, SB, IRBBB, and TWI are often benign in athletes. The PhysioNet Challenge weight matrix *W*_*ij*_ (used in the loss function, see supplementary Appendix F addresses this to some extent through weighted penalties. However, the matrix’s risk stratification is tuned for the general rather than the athlete population. Future work should incorporate an athlete-specific risk-stratified approach.

## Conclusion

This feasibility study demonstrates successful domain adaptation by classifying sports-related arrhythmias using models trained on general-population ECGs. We systematically analysed two model architectures using a novel xAI pipeline framework comprising data explainability, model interpretability, post-hoc explainability, and explainability assessment.

Regular and model-interpretable sinc convolution models achieve satisfactory classification of NSR, SB, IRBBB, and TWI, with distinct performance patterns. The sinc convolution network shows superior NSR detection (AUROC 0.75 vs 0.70), while regular convolution achieves better SB (AUROC 0.74 vs 0.70), IRBBB (AUROC 0.66 vs 0.58) and TWI (AUROC 0.59 vs 0.54) results. We hypothesise that sinc convolution layers better capture rhythms with periodicity and regularity but struggle with complex morphological patterns.

Grad-CAM analysis as post-hoc explainability identified that zero-padding significantly influences model behaviour, with sinc convolution showing greater sensitivity to non-physiological artefacts. Despite this, the sinc model focuses on physiologically relevant ECG segments (the PR segment for NSR/SB and the T-wave for TWI). Band-pass filtering with learnable cut-off frequencies aligns behaviour with underlying physiological signals, potentially improving interpretability for frequency-driven arrhythmias.

Future work should explore dynamic time warping, global attention mechanisms, class imbalance solutions, and population-specific datasets to enhance clinical applicability in distinguishing physiological athletic adaptations from pathological conditions.

## Data availability

The PhysioNet Challenge 21 and PF12RED datasets are open-source datasets and can be downloaded from https://physionet.org/content/challenge-2021/1.0.3/#files and https://github.com/dradolfomunoz/PF12RED respectively.

## Author contributions statement

EV: Conceptualisation, methodology, software, formal analysis, investigation, data curation, visualisation, writing—original draft. MH, PL: Clinical expertise, writing—review & editing. MV, LH, AB: Supervision, writing—review & editing.

