## Supplemental appendix for "An interpretable and explainable neural network to classify sports-related cardiac arrhythmias in professional football athletes"

The authors have provided this appendix to give readers additional information about their work.

**Funding information**      EV, LH, and MV are funded by The Podium Institute of Sports Medicine and Technology. AB is supported by the Royal Society University Research Fellowship (Grant No. URFR1‘314). PL acknowledges funding from the Oxford NIHR Biomedical Research Centre, Medical Research Council and British Heart Foundation. MH is funded by the National Institute for Health and Care Research (NIHR) Oxford Biomedical Research Centre (BRC). The views expressed are those of the authors and not necessarily those of the NIHR or the Department of Health and Social Care.

Appendix A. Exercise-induced cardiac remodelling in athletes

Exercise is a planned, structured, and repetitive physical activity to improve cardiorespiratory fitness (CRF) and reduce cardiovascular disease (CVD) risk and clinical outcomes in patients with known CVD ^1^. Physical activity involves repetitive energy expenditure above the resting metabolic rate and has cardioprotective effects ^2-4^.

Most arrhythmias on the pitch are due to undetected structural cardiac defects or channelopathies ^5^. These cardiac defects resemble the effects of exercise-induced cardiac remodelling (EICR), which are physiological and anatomical exercise-induced adaptations, such as cardiac chamber size and wall thickness increase due to exercise-induced volumes and pressure loads ^6^. Exercise-induced cardiac remodelling involves molecular, cellular, structural, and functional adaptations in response to repeated, elevated haemodynamic stress (see Fig. S1A). Myocardial hypertrophy for increased contraction and chamber dilatation to accommodate a higher blood volume are examples of structural remodelling ^7^. Higher ejection fraction to sustain the higher oxygen demand during exertion, and increased vagal tone and sympathetic stimulation while resting and exercising are functional- and electrical-induced adaptations, respectively ^8^.

Also known as the “athlete’s heart”, EICR effects involve and evolve into an ambiguous area between pathological and physiological cardiac adaptations. The prevailing assumption (founded on the preponderance of cardiovascular disorders) is that up to one out of 300 athletes may have an arrhythmic substrate for sudden cardiac arrest (SCA) predisposition ^9^. We define SCA as the precursor of SCD, in which the heart rhythm suddenly changes to a rhythm that could precipitate SCD, such as ventricular tachycardia.

Genetic variants known to predispose to arrhythmias (e.g. hypertrophic cardiomyopathy or arrhythmogenic cardiomyopathy) further increase an individual’s risk of life-threatening arrhythmias. However, these genetic variants are typically only tested for when there’s a concrete clinical suspicion or a known family history of the condition. Furthermore, some individuals develop the disease phenotype without carrying any well-known gene mutations, presumably because they have fewer common variants or mutations that are not commonly tested. Further acute modulating factors of the athlete’s arrhythmogenic phenomenon are haemodynamic stress, electrolytic changes, body temperature, and the environment ^8^.

The sport’s inherent statical (i.e., resistance) and dynamical (i.e., endurance) components also impact the exercise-induced cardiac remodelling. The main impact factors are intensity, duration, and frequency. The Morganroth hypothesis describes the relationship between the type of exercise (static or dynamic) and subsequent cardiac remodelling. Endurance training promotes eccentric ventricular remodelling, increasing the heart’s chamber volume and overall mass while conserving the wall thickness ^6,10^. Nevertheless, less evidence exists for the Morganroth hypothesis that resistance training results in concentric ventricular remodelling with increased wall and cardiac mass but minimal volume changes ^6,10^. This lack of clarity raises the question of whether resistance exercise is a distinct entity with its own mechanism or simply a less effective variant of endurance exercise ^16^. The Morganroth hypothesis was formulated solely on male participants, raising questions about whether it applies to the female sex. Regardless of sex, the heart adapts by increasing cardiac chamber size and wall thickness in response to the volume and pressure loads of exercise ^10^.

Huttin et al. ^11^ studied exercise-induced ECG changes in 2*,*484 French professional football players. The serial evaluations performed between 2005 and 2015 include 6*,*247 12-lead ECGs. The ECGs were recorded in the supine position during quiet respiration. A trained cardiologist evaluated the ECGs according to the European Society of Cardiology and Seattle criteria. The main results are summarised in Fig. S1B in the “ECG changes in football players” section. Sinus bradycardia (SB) at 98%, first-degree atrioventricular block (IAVB) at 31%, and short PR Interval at 80% (including incomplete right bundle branch block (IRBB)) are the main changes observed in the cohort. All these changes are part of training-related ECG changes, as listed in ^12^ (see section “ECG changes in athletes” in Fig. S1B).

The “extreme exercise hypothesis” outlines how the beneficial effects of sport may stagnate or decline when athletes exceed the optimal exercise dose ^13^. *The main question is how significant the sports-arrhythmia relationship is in each case, not whether one exists ^14^*. The interdependence of structural and electrical cardiac remodelling bedevils the mechanisms underlying the link between the athlete’s heart and arrhythmias. According to Heidbuchel ^14^, the following three elements contemplate the relation between sports and arrhythmia:

- *Sport triggers an arrhythmia on top of an underlying substrate*. This element emphasises that an athlete with an inherited or acquired structural or electrical pathology may develop a sports-related arrhythmia triggered by physical activity.
- *Sport facilitates an arrhythmic substrate or promotes* an underlying substrate development by accelerating the phenotype formation of arrhythmic events.
- *Sport yields a substrate*. This last element highlights that exercise-induced cardiac remodelling inherently creates a substrate for arrhythmias.

Thus, a limit in which exercise is not beneficial, on a long or even short-term basis, is also evident. Discerning between exercise-induced cardiac adaptations and cardiac pathologies with the potential for SCD is a fundamental clinical problem in sports medicine. Therefore, identifying the moment when the exercise-benefit ratio inverts and performance-enhancing adaptations may develop in adverse directions is critical ^15^. For this identification, a technology to classify cardiac rhythms in young athletes and detect arrhythmias is pivotal.

Appendix B. ECG data pre-processing

We first perform a lead expansion of ECG data. Expanding the Holter ECGs (i.e., 3-lead configuration) to a 12-lead configuration allows the data to be structured as a consistent 2D matrix, making it compatible with Conv2D layers. The expansion enables the model to effectively learn spatial patterns across leads, leveraging convolutional filters to extract meaningful features from ECG signals.

All ECGs are resampled to 500Hz. The resampling is needed because all but two datasets (INCART and PTB; see Table 6 in the Appendix) in the PhysioNet dataset are above or below 500Hz. A zero-phase 3rd-order Butterworth bandpass filter with a frequency range of 1Hz to 47Hz is used to filter the ECG data. This filter choice preserves the ECG waveform’s morphology while removing baseline wander (below 1Hz) and powerline interference (above 47Hz). We chose 47Hz because the data comes from geographically distinct locations worldwide. Depending on the region, the power supply’s frequency is 50Hz or 60Hz and harmonics ^16,17^.

A z-score normalises each of the samples of an ECG channel, ensuring that all ECG channels have a mean of 0 and a standard deviation of 1. The normalisation prevents features with larger absolute values (e.g., due to differences in amplitude across leads) from disproportionately influencing the neural network’s feature extraction. Then, the ECG data is filled with zeros in the time domain until reaching 8*,*192 samples. We chose 8*,*192 because it is the next power of 2, so the CPSC and CPSC-Extra (500Hz x 15*.*9s = 7*,*950) ECG signals can be allocated (see Table S3, column mean duration). Random sampling is performed if the signal is longer than 8,192 samples (INCART has 462,600 samples and PTB 110,800).

Appendix C. Model interpretability: The sinc convolution layer

Let $x[n]$ be the discrete input signal,$h[n]$ be the discrete filter, and $y[n]$ be the output signal. The standard convolution layer is defined by:

$y\left[ n \right]=x\left[ n \right]*h\left[ n \right]$

(1)

i.e., a convolution between the n-th sample of the ECG signal$x[n]$ and some finite impulse response filter $h[n]$. The SincNet proposed by Ravanelli and Bengio ^18^ filters the signal with a predefined function $g[n, \theta]$.

$$y\left[ n \right]=x\left[ n \right]*g\left[ n,\theta\right]$$

(2)

where θ is the parameter to learn. The function g only depends on the filter’s learnable parameter $\theta$, which controls the characteristics of the filter. In biological signals like ECGs, specific frequency ranges contain critical information for arrhythmia detection. A bandpass filter helps the network focus on the relevant frequency range, removing noise and irrelevant components, allowing the model to learn the optimal frequency bands that capture the key features of the ECG signal. Therefore, a filter bank of rectangular bandpass filters is a reasonable choice for $g$ ^18^, since bandpass filters allow frequencies within a specific range and attenuate all others outside that range. Equation (3) defines a bandpass filter in the frequency domain as:

$$G\left( f, f_{1}, f_{2} \right)=rect\left( \frac{f}{2f_{2}} \right)- rect\left( \frac{f}{2f_{1}} \right)$$

(3)

where the parameters $f_{1}$ and $f_{2}$ are the filter’s low and high cut-off frequencies, respectively, and $rect()$ is the rectangular bandpass filter function defined as:

$$\mathrm{rect}\left( x \right)=\left\{ \begin{aligned} 0, if f< f_{1} \\ 1, if f_{1}< f<f_{2} \\ 0, if f \geq f_{2} \end{aligned} \right.$$

(4)

$G(f, f_{1}, f_{2})$ represents the output of the filter in the frequency domain, and $f$ represents the frequency at which we evaluate the filter’s behaviour. The filter passes the frequency $f$ if it lies between $f_{1}$ and $f_{2}$. Through an inverse Fourier transformation, the bandpass in the time domain is defined as the following equation:

$$g\left[ n,f_{1},f_{2} \right]=2f_{2}\mathrm{sinc}\left( 2\pi f_{2}n \right)-2f_{1}\mathrm{sinc}\left( 2\pi f_{1}n \right)$$

(5)

Inserting Eq. (5) into Eq. (2) results in the following expected filtered output $y[n]$ :

$$y\left[ n \right]=x\left[ n \right]*2f_{2}\mathrm{sinc}\left( 2\pi f_{2}n \right)-2f_{1}\mathrm{sinc}\left( 2\pi f_{1}n \right)$$

(6)

Comparing Eq. (2) with Eq. (6) clarifies that the cut-off frequencies $f_{1}$ and $f_{2}$ correspond to the learnable parameter $\theta$. This approach enables a trade-off between leveraging full convolutional potential in subsequent layers and ensuring interpretability by passing information with physical meaning through the first convolutional layer. The interpretability is due to the learnable parameters θ in the first layer of the network, which are the cut-off frequencies $f_{1}$ and $f_{2}$. The parameters $f_{1}$ and $f_{2}$ directly relate to the signal's frequency range, determining the bandwidth the model focuses on. The neural network inherently incorporates domain-specific knowledge with physical meaning by constraining its weights to specific frequency ranges. This constraint provides transparency into the feature extraction process, rather than learning arbitrary features across the entire frequency spectrum.

Appendix D. Downsampling the feature maps: pooling layer

A dropout layer drops half of the neurons to prevent overfitting. Then, the dropout layer’s output runs through the multi-head mechanism, with eight heads on the 256-channel data, meaning each head focuses on 32 dimensions. The multi-head’s attention mechanism results are converted into a 256-dimensional vector. Further, an adaptive max pooling layer reduces the dimensionality by downsampling each of the 256 feature maps to a single maximum value.

Appendix E. Classifying the arrhythmias: the fully connected layer

The fully connected layer combines the attention mechanism and pooling output with the lead indicator and outputs two prediction heads. The first output head contains a binary cross-entropy (BCE) loss function. In contrast, the second output forms an additional neural network (no gradient propagation) with a challenge (CL) and sparsity loss function (SL) (see Appendix for an explanation of CL and SL). The total loss function is defined as:

$$Loss =\sum_{\mathrm{batch}} \mathrm{BCE}\left( t,p \right)-\mathrm{CL}\left( t,p \right)+\mathrm{SL}\left( p \right)$$

(7)

with *p* being the probabilities and *t* being the targets.

The modified confusion matrix with entries defines the CL $a_{ij}$ and corresponding weights $w_{ij}$ given by the PhysioNet ^19^, resulting in:

$$\mathrm{CL}\left( t, p \right)=\sum_{\mathrm{batch}} w_{ij}a_{ij}\left( t,p \right)$$

(8)

The SL derived from the parabolic curve is defined as $SL(p) = -4p(p - 1$), encouraging the model to output probabilities close to 0 or 1 to make definitive decisions regarding arrhythmias rather than being uncertain ^20^. An Adam optimiser (epoch 30 with learning rate 1 × 10−3) trains the model with a batch size of 128 and an L2 regularisation parameter of 1 × 10−4. At the 20th epoch, the learning rate is reduced by a factor of 0.10.

Appendix F. Post-hoc explainability: Grad-CAM

First, the Grad-CAM performs a forward pass to get the specific ECG's target class $c$ (i.e., cardiac rhythm) prediction. Then, the Grad-CAM computes the gradient of the score $y^{c}$ of the target class *c* concerning feature map activations $A^{k}$ of a chosen convolutional layer, i.e., $\frac{\partial y^{c}}{\partial A^{k}}, k\in\left\{ 1,2,\ldots,256 \right\}$. In this context, $y^{c}$ represents the logit for a specific cardiac rhythm such as IRBBB or TWI (i.e., pre-softmax output). Grad-CAM uses these pre-softmax scores because these scores are not compressed into the 0 to 1 range and, therefore, represent the network’s “reasoning” before normalisation.

Usually, the chosen convolutional layer is the last layer, where the feature maps $A^{k}$ represent the patterns detected by the last layer. The derivative $\frac{\partial y^{c}}{\partial A^{k}}$calculates how a slight change in each feature map $A^{k}$ would affect the prediction for class $c$.

The gradients $\frac{\partial y^{c}}{\partial A^{k}}$are globally averaged and pooled to obtain weights $\alpha^{c}$ that represent the importance of each feature map for the target class. In this context, global average pooling determines which ECG feature map (e.g., P-wave, QRS-complex, T-wave) is most important for identifying the target class $c$. The Grad-CAM results in:

$$a_{k}^{c}= \frac{1}{Z}\sum_{i} \sum_{j} \frac{\partial y^{c}}{\partial A^{k}}$$

(9)

where in the ECG context, $Z$ is a normalisation factor, and$i$ and $j$ feature maps’ spatial dimension, time and ECG lead. To create the actual heatmap, the weighted sum $L^{c}=\sum_{k} \alpha_{k}^{c}A^{k}$ is created. Because the weighted sum $L^{c}$ contains both positive and negative influences, a ReLU function is applied to only show ECG segments that strongly influence the classification, i.e.:

$L_{Grad-CAM}^{c}=ReLU\left( \sum_{k} \alpha_{k}^{c}A^{k} \right)$

(10)

Lastly, the resulting heatmap $L_{Grad-CAM}^{c}$ isupsampled from the output size of the chosen convolutional layer to the input size of the ECG input signal (i.e., 8,192). We saved 15 Grad-CAM ECG examples for each cardiac rhythm.

Appendix G. Cross-validation

Before testing on the sports dataset, baseline performance was established using 3-fold cross-validation on the PhysioNet dataset with regular convolution layers (see Fig. S3). Each fold maintained the same arrhythmia distribution as the full dataset (deviation in arrhythmia distribution between folds <1%), providing a methodical baseline for comparison when training, validation, and testing were used on the same dataset.

The 3-CV baseline results are summarised in Table S1. All CVs achieved AUROC performance between 0.85 and 0.87, AUPRC between 0.28 and 0.29, and F1-scores between 0.28 and 0.32. Challenge scores ranged from 0.57 to 0.59 for 30-epoch training. We do not include the challenge score in subsequent results, since calculating it based on only four rhythms (NSR, sinus bradycardia (SB), IRBBB, and TWI) would yield a poor, non-representative score. The best CV was CV2 (see Table S2). Results for the four shared cardiac rhythms ranged between 0.82–0.97, 0.1–0.83, and 0.2–0.84 for AUROC, AUPRC, and F1-score, respectively.

Appendix H. Tables
Table S1 3-CV baseline results (AUROC, AUPRC, F1-score, and Challenge score) on the PhysioNet Challenge 21 dataset. CV 2 outperformed CV 1 and CV 3 in all three metrics. We included the results for CV 2 regarding the cardiac rhythms present in the PF12RED dataset in Table S2.

| CV | AUROC | AUPRC | F1-score | Challenge score |
| --- | --- | --- | --- | --- |
| CV 1 | 0*.*85 | 0*.*28 | 0*.*31 | 0*.*57 |
| CV 2 | 0*.*87 | 0*.*29 | 0*.*32 | 0*.*59 |
| CV 3 | 0*.*85 | 0*.*28 | 0*.*28 | 0*.*59 |

Abbreviations: CV denotes cross-validation.

Table S2 Results of CV 2 (see Table S1) on the PhysioNet Challenge 21 dataset for the four cardiac rhythms are also contained in the PF12RED dataset.

| Label | AUROC | AUPRC | F1-score |
| --- | --- | --- | --- |
| NSR | 0*.*85 | 0*.*69 | 0*.*72 |
| SB | 0*.*97 | 0*.*83 | 0*.*84 |
| IRBBB | 0*.*83 | 0*.*10 | 0*.*20 |
| TWI | 0*.*82 | 0*.*15 | 0*.*25 |

| Dataset | ECG | Sampling frequency (Hz) | Mean  duration  (s) | Labelling procedure | ECG evaluation criteria |
| --- | --- | --- | --- | --- | --- |
| Chapman-Shaoxing | 12-leads | 500 | 10 | Two physicians individually, in case of disagreement, a third, senior physician adjudicated | — |
| CPSC | 12-leads | 500 | 15.9 | — | — |
| CPSC-Extra | 12-leads | 500 | 15.9 | — | — |
| G12EC | 12-leads | 500 | 10 | — | — |
| INCART | 12-leads | 257 | 1,800 | Diagnosis confirmed by enzyme assays, coronary angiography,  electrophysiological study, and pressure monitoring where needed | — |
| Ningbo | 12-leads | 500 | 10 | Labelled by a cardiologist-supervised  physician | — |
| PTB | 12-leads | 1000 | 110.8 | Medical findings and diagnoses  confirmed by several examinations | — |
| PTB-XL | 12-leads | 500 | 10 | Up to two cardiologists | — |
| PF12RED | 12-leads | 500 | 10 | One  cardiologist | International  Criteria for  Electrocardiogram  Interpretation  in Athletes |

Table S3 Technical information, like the recording device, sampling frequency, mean recording duration and used labelling procedure regarding the ECG recordings of every single PhysioNet Challenge 21 dataset ^21^ and PF12RED dataset ^22^. Five sources do not mention any recording device.
Appendix I. Figures

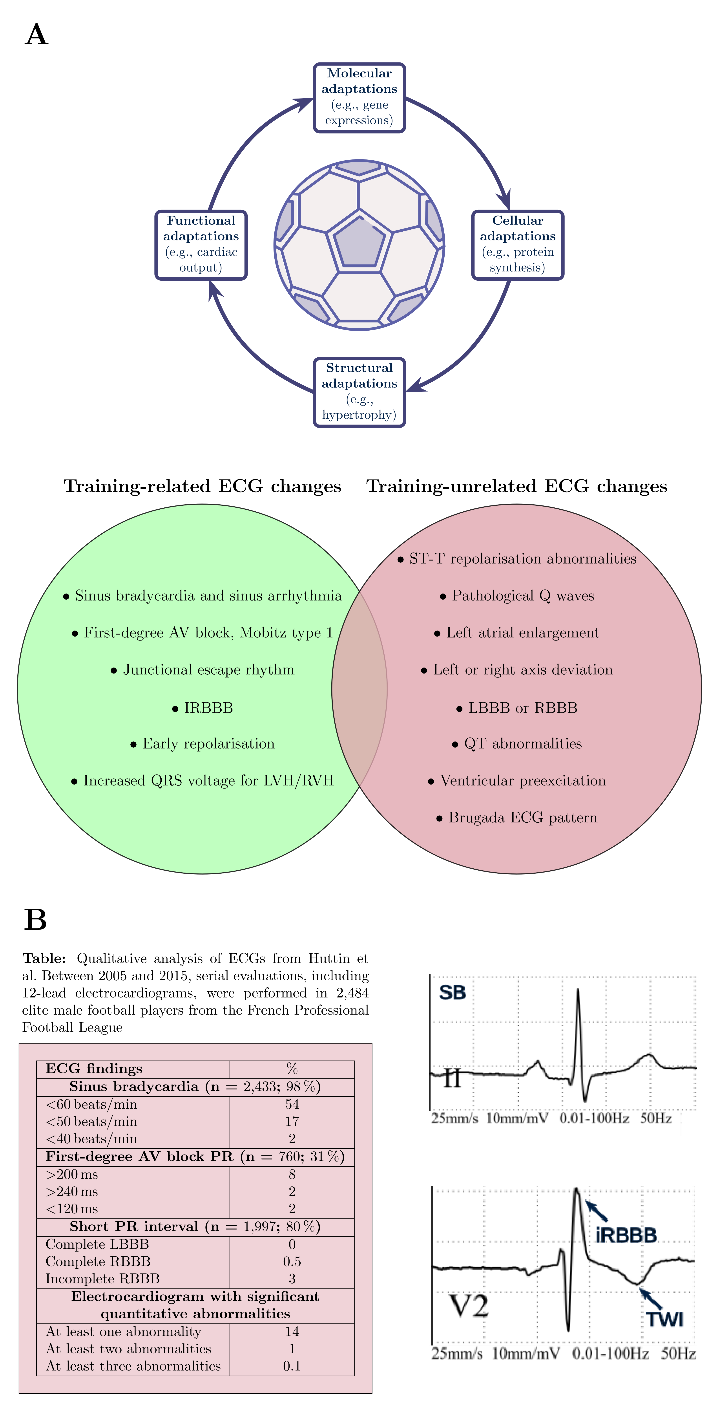

Figure S1 **Exercise-induced cardiac remodelling and ECG changes in athletes and footballers. A)** Exercise-induced cardiac adaptations include molecular, cellular, structural, and functional adaptations. The ECG displays these adaptations, which can be training-related or training-unrelated. **B)** ECG changes in football athletes from Huttin et al ^11^. In a clinical study, French professional football players showed similar training-related ECG changes, with SB being the most notable, at 98%. On the contrary, 14% had training-unrelated ECG changes or abnormalities which required further investigation. AV denotes atrioventricular; ECG, electrocardiogram; IRBBB, incomplete right bundle branch block; LBBB, left bundle branch block; RBBB, right bundle branch block; and TWI, T-wave inversion.

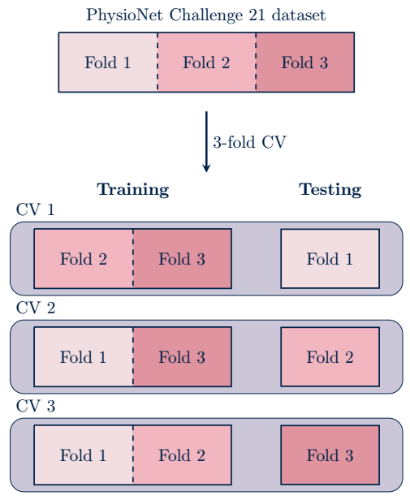

Figure S2 **Visualisation of the 3-fold CV.** 3-fold CV for training, validation, and testing of the neural network with a regular convolution layer on the PhysioNet Challenge 21 dataset. The deviation of the three folds is below 1% compared to the entire dataset. CV denotes cross-validation.

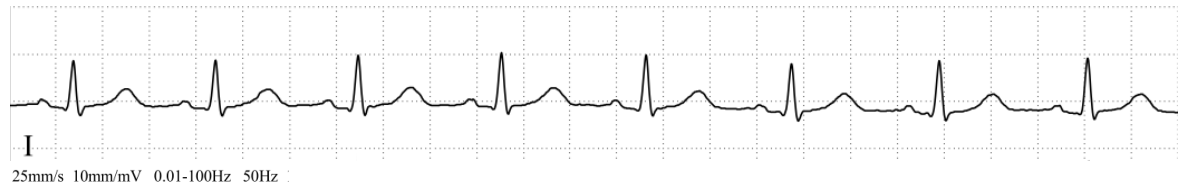

*(a)*

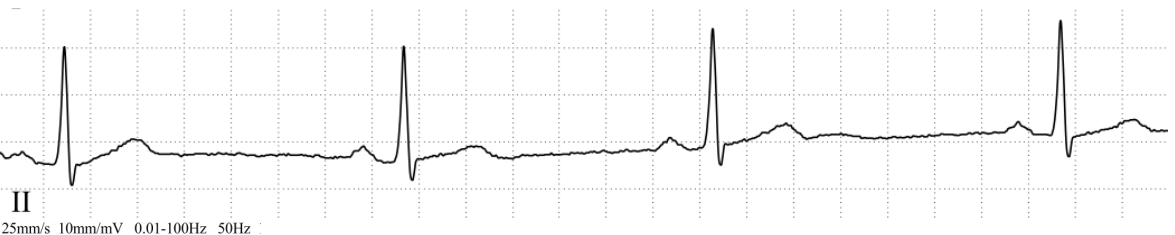

*(b)*

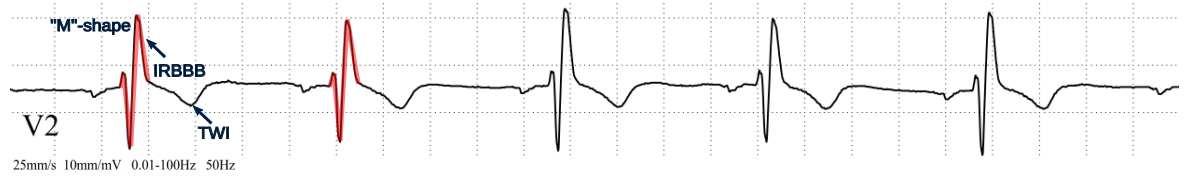

*(c)*

Figure S3 **Examples of cardiac rhythms from the PF12RED dataset.** 5 s of the cardiac rhythms present in the PF12RED dataset: (a) NSR (player ID 36), (b) SB (player ID 19), (c) IRBBB and TWI (player ID 33). While the inversion of the T-wave can recognise the TWI, the rSR’ pattern is characteristic for the IRBBB, here appearing as the letter “M” (highlighted in red on the first two QRS-complexes). NSR denotes normal sinus rhythm; SB, sinus bradycardia; IRBBB, incomplete right bundle branch block; and TWI, T-wave inversion.
